# Evaluating the effect of public health intervention on the global-wide spread trajectory of Covid-19

**DOI:** 10.1101/2020.03.11.20033639

**Authors:** Zixin Hu, Qiyang Ge, Shudi Li, Li Jin, Momiao Xiong

## Abstract

As COVID-19 evolves rapidly, the issues the governments of affected countries facing are whether and when to take public health interventions and what levels of strictness of these interventions should be, as well as when the COVID-19 spread reaches the stopping point after interventions are taken. To help governments with policy-making, we developed modified auto-encoders (MAE) method to forecast spread trajectory of Covid-19 of countries affected, under different levels and timing of intervention strategies. Our analysis showed public health interventions should be executed as soon as possible. Delaying intervention 4 weeks after March 8, 2020 would cause the maximum number of cumulative cases of death increase from 7,174 to 133,608 and the ending points of the epidemic postponed from Jun 25 to Aug 22.

---

As of March 8, 2020, global confirmed cases of COVID-19 passed 105,587 and has spread to 102 countries. The coronavirus is approaching dangerous new stages (**1**). Is the spread of coronavirus unstoppable? Is a COVID-19 pandemic inevitable. There is an urgent need to develop strategies to control the spread of COVID-19.

A number of the statistical, dynamic and mathematical models of the Covid-19 outbreak including the SEIR model, branching processes have been developed to analyze its transmission dynamics (2-7). Although these epidemiological models are useful for estimating the dynamics of transmission, targeting resources and evaluating the impact of intervention strategies, the models require parameters and depend on many assumptions. Unlike system identification in engineering where the parameters in the models are estimated using real data, at the outbreak, estimated parameters using real time data are not readily available (8-9).

Most analyses used hypothesized parameters and hence do not fit the data very well. The accuracy of forecasting the future cases of Covid-19 using these models may not be very high. The intervention strategies that are developed by these models cannot be evaluated by real data. Timely interventions are needed to control the serious impacts of Covid-19 on health.

To overcome limitations of the epidemiological model approach and assist public health planning and policy making, we developed the modified auto-encoder (MAE), an artificial intelligence (AI) based method for real time forecasting of the new and cumulative confirmed cases of Covid-19 under various interventions in more than 100 countries across the world (10,11). The MAE can model interventions, while still using real data for evaluation of interventions. Transfer learning was used to train the MAE (12). An intervention variable was introduced as an input variable for the MAE. We viewed the China type of intervention as the fully comprehensive intervention and assigned 1 to the intervention variable. We assigned 0 to the intervention variable if there was no intervention. The weights between 0 and 1 were assigned to the intervention variable for the different degrees of interventions. The values that were assigned to the intervention variable was called weight. Taking time for intervention into account, we considered four comprehensive intervention scenarios. The first intervention scenario was to start intervention on March 9, 2020 and the weight was 1. The second intervention scenario was to start limited intervention on March 9 (the weight was 0.5) and one week later to start complete intervention (the weight was 1). The third intervention scenario was that in the first week (starting on March 9) no intervention was taken (the weight was 0); in the second week, take limited intervention (the weight was 0.5); and after the third week, take full intervention (the weight was 1). The fourth intervention scenario was that in the first one week, no intervention was taken (the weight was 0), in the next three weeks, limited intervention was taken (the weight was 0.5), and after the fourth week, full intervention was taken (the weight was 1). We investigate how the degree of intervention and starting intervention time determine the peak time and case ending time, the peak number and maximum number of cases and forecasting the peak and maximum number of new and cumulative cases in more than 100 countries across the world. The analysis is based on the surveillance data of confirmed and new Covid-19 cases in the world up to March 8, 2020.

## Later intervention makes it difficult to stop the spread of COVID-19

Data on the number of confirmed, new and death cases of Covid-19 from January 20, 2020 to March 8 were from WHO (https://www.who.int/emergencies/diseases/novel-coronavirus-2019/situation-reports). Data included the total numbers of the accumulated, newly confirmed and death cases in the world and across 102 countries.

To demonstrate that the modified auto-encoder (MAE) is a reasonable forecasting method, the MAE was applied to the confirmed accumulated cases of Covid-19 across 102 countries in the world. The intervention indicator for China and other countries was set to 1 and 0, respectively. Table S1 presented the one-step to five-step errors for forecasting the accumulated cases, starting from March 3, 2020. Table S1 showed that in any cases the forecasting accuracies of the MAE were less than 5%.

We consider comprehensive interventions. Comprehensive intervention included strict travel restriction, cancel conferences, mandatory quarantine, restrict public transportation, school closing and shut down all non-essential companies. Using real data to evaluate the consequences of specific intervention is infeasible. We considered three intervention scenarios: (1) China type intervention where 1 was assigned to the intervention variable, (2) no major interventions where 0 was assigned to the intervention variable and (3) between the China type intervention and no major interventions where 0.5 was assigned to the intervention variable. Table 1 presented the forecasting results of COVID-19 in 30 countries and worldwide under limited and later intervention (scenario 4). Table 1 showed that the total peak number of confirmed cases and new cases in the world with limited intervention could reach 1,516,300 and 178,090, respectively. Table 1 also showed that if every country in the world took later intervention, finally, the total number of cases in the world could reach as high as 4 million, a frightening number (3,929,641) and miserable transmission of COVID-19 continuing until late Aug 2020. Table 1 also showed that the top 10 countries with a high average increasing rate of the cases were Italy, Iran, Germany, USA, South Korea, Netherlands, France, China and Switzerland. To show the dynamics of COVID-19, Figure 1(G) and (H) presented plotted the curves of the confirmed cases and new cases of seven major infected countries: China, Iran, Italy, Germany, South Korea, USA, France and all other countries under scenario 4. We observed that numbers of new cases in all countries except for China would peak in early April. Both sides of the peak of the new cases will be steep, which implies that the new cases will increase rapidly and put great pressure on the health care workers and facilities when the curve approaches the peak.

**Table 1.** Spread of COVID-19 in 30 countries and whole world under limited intervention.

| State | Peak Time | End Time | Duration | Peak (Cum) | Peak (New) | Current Case | End Case | Up Slope | Down Slope |
| --- | --- | --- | --- | --- | --- | --- | --- | --- | --- |
| Total | 2020/4/5 | 2020/8/22 | 215 | 1,516,300 | 178,090 | 105,587 | 3,929,641 | 2,160.90 | -1,200.18 |
| Italy | 2020/4/9 | 2020/8/8 | 191 | 781,177 | 70,705 | 5,883 | 1,493,498 | 973.22 | -521.66 |
| Iran | 2020/4/5 | 2020/8/22 | 185 | 317,757 | 32,808 | 5,823 | 729,274 | 662.98 | -224.18 |
| Germany | 2020/4/9 | 2020/7/28 | 183 | 153,230 | 13,960 | 795 | 293,797 | 183.63 | -112.75 |
| USA | 2020/4/9 | 2020/7/17 | 177 | 75,068 | 6,890 | 213 | 144,542 | 84.53 | -61.55 |
| Korea | 2020/4/5 | 2020/7/25 | 187 | 57,544 | 5,071 | 7,134 | 121,663 | 63.61 | -44.13 |
| Netherlands | 2020/4/9 | 2020/7/18 | 142 | 59,325 | 5,400 | 188 | 113,773 | 127.64 | -47.87 |
| France | 2020/4/5 | 2020/7/29 | 187 | 40,788 | 4,185 | 706 | 93,277 | 54.47 | -34.59 |
| Belgium | 2020/4/9 | 2020/7/13 | 160 | 44,870 | 4,039 | 169 | 85,461 | 60.31 | -37.77 |
| China | 2020/2/5 | 2020/5/19 | 130 | 31,432 | 5,236 | 80,859 | 82,740 | NA | 0.00 |
| Switzerland | 2020/4/5 | 2020/8/2 | 159 | 35,657 | 3,714 | 264 | 82,431 | 86.71 | -29.87 |
| Sweden | 2020/4/5 | 2020/7/18 | 169 | 32,195 | 3,585 | 161 | 77,364 | 52.02 | -33.01 |
| UK | 2020/4/5 | 2020/7/18 | 169 | 25,070 | 3,360 | 210 | 71,762 | 47.44 | -30.12 |
| Austria | 2020/4/9 | 2020/7/26 | 152 | 27,241 | 2,458 | 104 | 51,954 | 55.39 | -20.29 |
| Lebanon | 2020/4/9 | 2020/7/5 | 135 | 20,797 | 1,909 | 28 | 39,998 | 38.98 | -19.27 |
| Spain | 2020/4/5 | 2020/7/9 | 160 | 16,260 | 1,617 | 430 | 36,408 | 23.11 | -16.05 |
| Greece | 2020/4/9 | 2020/7/1 | 126 | 18,223 | 1,672 | 66 | 35,061 | 38.48 | -17.69 |
| Canada | 2020/4/9 | 2020/7/1 | 157 | 16,674 | 1,525 | 57 | 31,988 | 19.76 | -16.13 |
| Australia | 2020/4/9 | 2020/7/5 | 163 | 16,360 | 1,494 | 74 | 31,406 | 18.85 | -15.14 |
| Japan | 2020/4/5 | 2020/7/20 | 182 | 12,652 | 1,257 | 455 | 28,449 | 15.53 | -11.28 |
| Malaysia | 2020/4/5 | 2020/7/11 | 168 | 10,228 | 1,074 | 93 | 23,776 | 14.21 | -10.57 |
| UAE | 2020/4/5 | 2020/7/9 | 163 | 9,639 | 1,037 | 45 | 22,514 | 14.29 | -10.41 |
| Singapore | 2020/4/9 | 2020/6/11 | 140 | 11,268 | 1,011 | 138 | 21,390 | 12.61 | -13.93 |
| Portugal | 2020/4/9 | 2020/6/28 | 118 | 9,592 | 877 | 21 | 18,388 | 23.03 | -9.61 |
| Brazil | 2020/4/9 | 2020/6/10 | 105 | 8,955 | 815 | 19 | 17,120 | 18.80 | -11.39 |
| Ireland | 2020/4/9 | 2020/6/23 | 115 | 8,024 | 737 | 19 | 15,384 | 18.27 | -8.56 |
| Norway | 2020/4/5 | 2020/7/1 | 126 | 6,646 | 663 | 147 | 14,904 | 15.75 | -7.24 |
| Iceland | 2020/4/9 | 2020/6/19 | 110 | 7,469 | 690 | 45 | 14,368 | 17.56 | -8.44 |
| Finland | 2020/4/9 | 2020/6/28 | 151 | 7,333 | 698 | 19 | 14,360 | 9.31 | -7.54 |
| Demark | 2020/4/9 | 2020/6/26 | 121 | 6,741 | 614 | 31 | 12,887 | 14.13 | -6.89 |
| Kuwait | 2020/4/9 | 2020/6/10 | 108 | 5,876 | 534 | 62 | 11,227 | 11.40 | -7.45 |

**Fig. 1.**
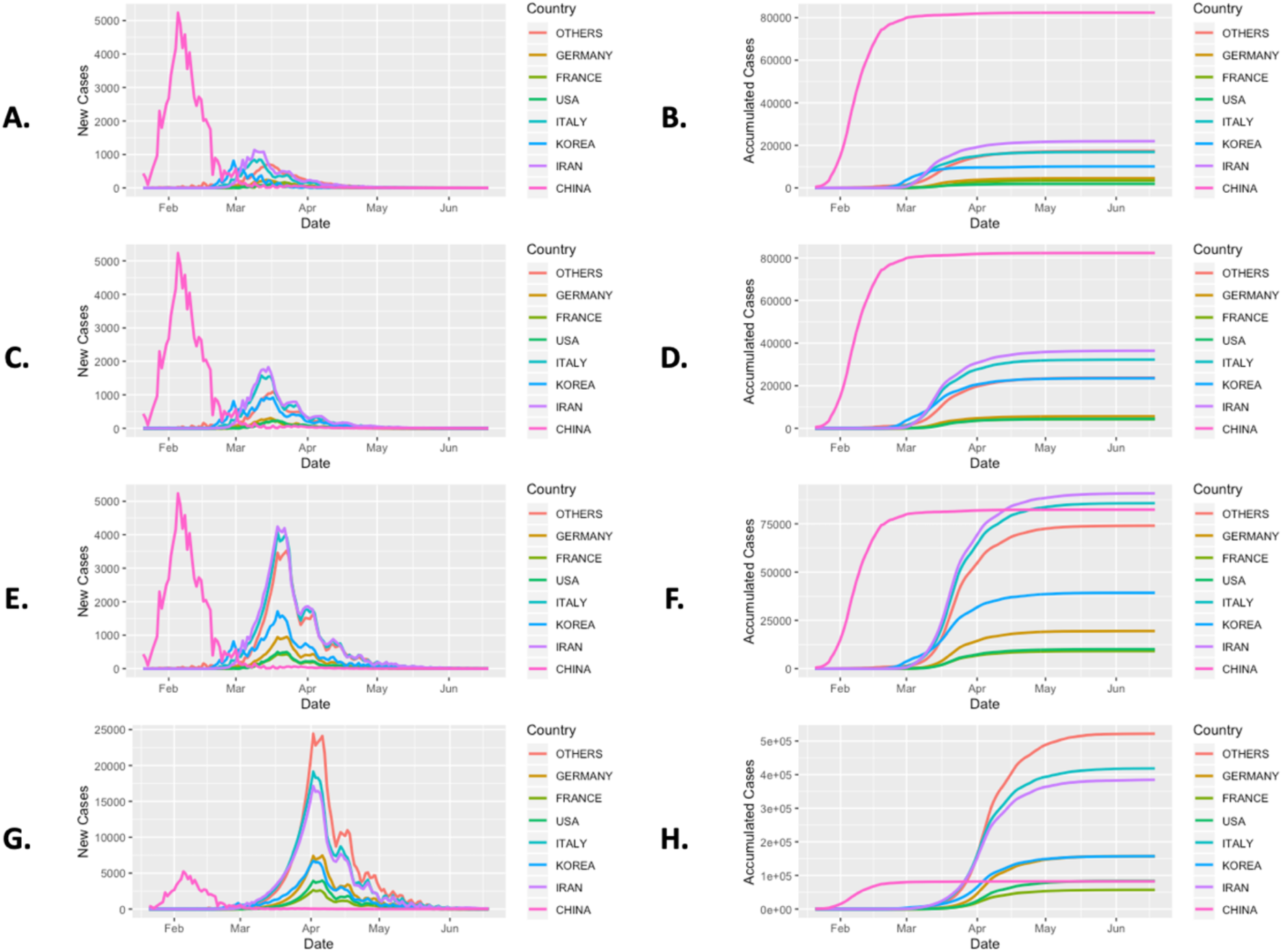
Trajectory of COVID-19 in seven most infected countries: China, Iran, Italy, Germany, South Korea, USA, France, and all other countries as a function of days from January 21 to June 18, 2020. (A), (C), (E) and (G) Forecasted curves of the newly confirmed cases of Covid-19 under scenarios 1, 2, 3 and 4, respectively.(B), (D), (F) and (H) Forecasted curves of the cumulative confirmed cases of Covid-19 under scenarios 1, 2, 3 and 4, respectively.

## New strategies are needed to curb the spread of COVID-19

When we are past the point of containing coronavirus, there is urgent need to develop new strategies to curb the spread of COVID-19 (1). In this section, we investigate whether comprehensive interventions can control the spread of COVID-19 and how interventions will reduce the peak time and the number, and the final total number of cases across the world.

Table 2 showed the forecasted results of COVID-19 in 30 countries and world wide under immediate active intervention such as restrict social interaction, community mitigation measures and quarantine (Scenario 1). We can observe dramatic reduction of the cases of COVID-19. After taking active intervention, the forecasted final total number of cases (when the spread of COVID-19 is over) in the world was reduced from 3,929,641 to 211,000, i.e, 94.6% of the potential cases would be eliminated by active intervention, duration time was reduced from 215 days to 157 days, and the end time would change from Aug 22 to Jun 25. To show transmission changes of COVID-19, we presented Figure 1(A) and (B) that plotted the curves of the confirmed cases and new cases in seven major infected countries: China, Iran, Italy, Germany, South Korea, USA, France and all other countries under scenario 1. Figures showed that both new and cumulated cases of coronavirus outside China were much lower than that of China.

**Table 2.** Spread of COVID-19 in 30 countries and whole world under active intervention.

| State | Peak Time | End Time | Duration | Peak (Cum) | Peak (New) | Curr Case | End Case | Up Slop | Down Slop |
| --- | --- | --- | --- | --- | --- | --- | --- | --- | --- |
| Total | 2020/3/14 | 2020/6/25 | 157 | 133,312 | 5,436 | 105,587 | 211,000 | 72.04 | -36.39 |
| China | 2020/2/5 | 2020/5/19 | 130 | 31,432 | 5,236 | 80,859 | 82,740 | NA | 0 |
| Iran | 2020/3/14 | 2020/6/25 | 127 | 15,505 | 1,986 | 5,823 | 37,428 | 62.74 | -10.94 |
| Italy | 2020/3/14 | 2020/6/19 | 141 | 13,899 | 1,498 | 5,883 | 29,475 | 31.85 | -12.07 |
| Korea | 2020/2/29 | 2020/5/28 | 129 | 3,150 | 813 | 7,134 | 11,166 | 4.94 | -2.99 |
| USA | 2020/3/21 | 2020/6/12 | 142 | 3,444 | 456 | 213 | 8,795 | 1.98 | -0.97 |
| Germany | 2020/3/6 | 2020/5/24 | 118 | 534 | 272 | 795 | 5,278 | 3.4 | -1.86 |
| France | 2020/3/7 | 2020/5/26 | 123 | 613 | 193 | 706 | 3,095 | 2.82 | -1.63 |
| UK | 2020/3/17 | 2020/5/25 | 115 | 1,225 | 145 | 210 | 2,880 | 1.79 | -0.88 |
| Belgium | 2020/3/16 | 2020/5/21 | 107 | 1,076 | 136 | 169 | 2,595 | 2.24 | -1.03 |
| Sweden | 2020/3/23 | 2020/5/27 | 117 | 1,280 | 124 | 161 | 2,467 | 0.61 | -0.29 |
| Finland | 2020/3/29 | 2020/5/26 | 118 | 1,032 | 108 | 19 | 2,172 | 0.53 | -0.27 |
| Switzerland | 2020/3/7 | 2020/5/19 | 84 | 209 | 123 | 264 | 2,127 | 4.31 | -0.78 |
| Australia | 2020/3/22 | 2020/5/23 | 120 | 896 | 109 | 74 | 2,115 | 0.07 | -0.07 |
| Malaysia | 2020/3/21 | 2020/5/23 | 119 | 1,009 | 109 | 93 | 2,031 | 0.43 | -0.28 |
| Netherlands | 2020/3/15 | 2020/5/12 | 75 | 813 | 104 | 188 | 1,922 | 6.91 | -1.17 |
| Spain | 2020/3/7 | 2020/5/15 | 105 | 374 | 117 | 430 | 1,703 | 2.89 | -1.62 |
| Austria | 2020/3/17 | 2020/5/18 | 83 | 745 | 94 | 104 | 1,703 | 2.62 | -0.49 |
| Japan | 2020/3/12 | 2020/5/10 | 111 | 700 | 72 | 455 | 1,499 | 1.08 | -0.84 |
| UAE | 2020/3/21 | 2020/5/15 | 108 | 603 | 74 | 45 | 1,262 | 0.15 | -0.13 |
| Greece | 2020/3/24 | 2020/5/15 | 79 | 546 | 59 | 66 | 1,140 | 0.83 | -0.15 |
| Iceland | 2020/3/23 | 2020/5/8 | 68 | 398 | 43 | 45 | 805 | 0.5 | -0.08 |
| Norway | 2020/3/12 | 2020/5/1 | 65 | 301 | 44 | 147 | 730 | 2.92 | -0.65 |
| Singapore | 2020/3/15 | 2020/4/27 | 95 | 302 | 32 | 138 | 656 | 0.52 | -0.48 |
| Portugal | 2020/3/17 | 2020/5/3 | 62 | 211 | 31 | 21 | 532 | 0.71 | -0.11 |
| India | 2020/3/26 | 2020/5/8 | 100 | 290 | 25 | 34 | 506 | 0.15 | -0.1 |
| Brazil | 2020/3/16 | 2020/4/21 | 55 | 122 | 17 | 19 | 305 | 0.67 | -0.18 |
| Denmark | 2020/3/17 | 2020/4/25 | 59 | 146 | 17 | 31 | 305 | 0.5 | -0.13 |
| Canada | 2020/3/13 | 2020/4/22 | 87 | 121 | 20 | 57 | 302 | 0.19 | -0.18 |
| Lebanon | 2020/3/9 | 2020/4/21 | 60 | 48 | 20 | 28 | 271 | 1.12 | -0.43 |
| Romania | 2020/3/20 | 2020/4/23 | 57 | 117 | 13 | 13 | 225 | 0.17 | -0.04 |

It is highly unlikely to completely shut down cities. However, it is possible to use precision quarantine to slow down and keep people from transmitting the disease. To simulate the intervention measures between China with complete control and without any control, we presented Tables S2 and S3 which showed the results under scenarios 2 and 3. Figure 1(C), (D), (E) and (F) showed transmission dynamics of COVID-19 with plotted curves of the confirmed cases and new cases in the seven major infected countries: China, Iran, Italy, Germany, South Korea, USA, France and all other countries under scenarios 2 and 3, respectively. Scenarios 2 and 3 may be more realistic.

## Comparisons among several strategies

To further illustrate the impact of interventions on the spread of COVID-19, we comprehensively compare the effects of the four intervention scenarios on the transmission dynamics of COVID-19 across the world. Figure 2 plotted the world reported and forecasted time curves of the cumulative and newly confirmed cases of Covid-19 under four intervention scenarios. The ratios of the world number of final cases in the four scenarios were 1:1.5:3.3:18.6 and ratios of case duration under the four intervention scenarios were 1:1:1.2:1.4. These results showed that intervention time delays had serious consequences. Later intervention may increase almost 20 fold (18.6) the number of cases when Covid-19 would be over and will delay the ending time by almost two months. The ratio of the average increase index of new cases under the four intervention scenarios were respectively 1:2.88:5.88:30. Delaying intervention will substantially speed the spread of coronavirus. Figure S2 presented the forecasted duration, cumulative cases of outbreak end, and risk of the top 44 outbreak countries in the world where the risk index was defined as the ratio of the number of cumulative cases under intervention scenario *i, i* = 2,3,4 over the number of cumulative cases under intervention scenario 1. We observed that risks of later intervention among the top the 43 countries(China excluded) varied from 7 (Finland) to 241 (Ecuador). In other words, among these countries, the maximum number of cumulative cases under the later intervention scenario would increase at least 240 times more than that of early intervention. We also observed that the intervention strategy to control the spread of the coronavirus of the two countries: South Korea and Singapore were similar to that of China.

**Fig. 2.**
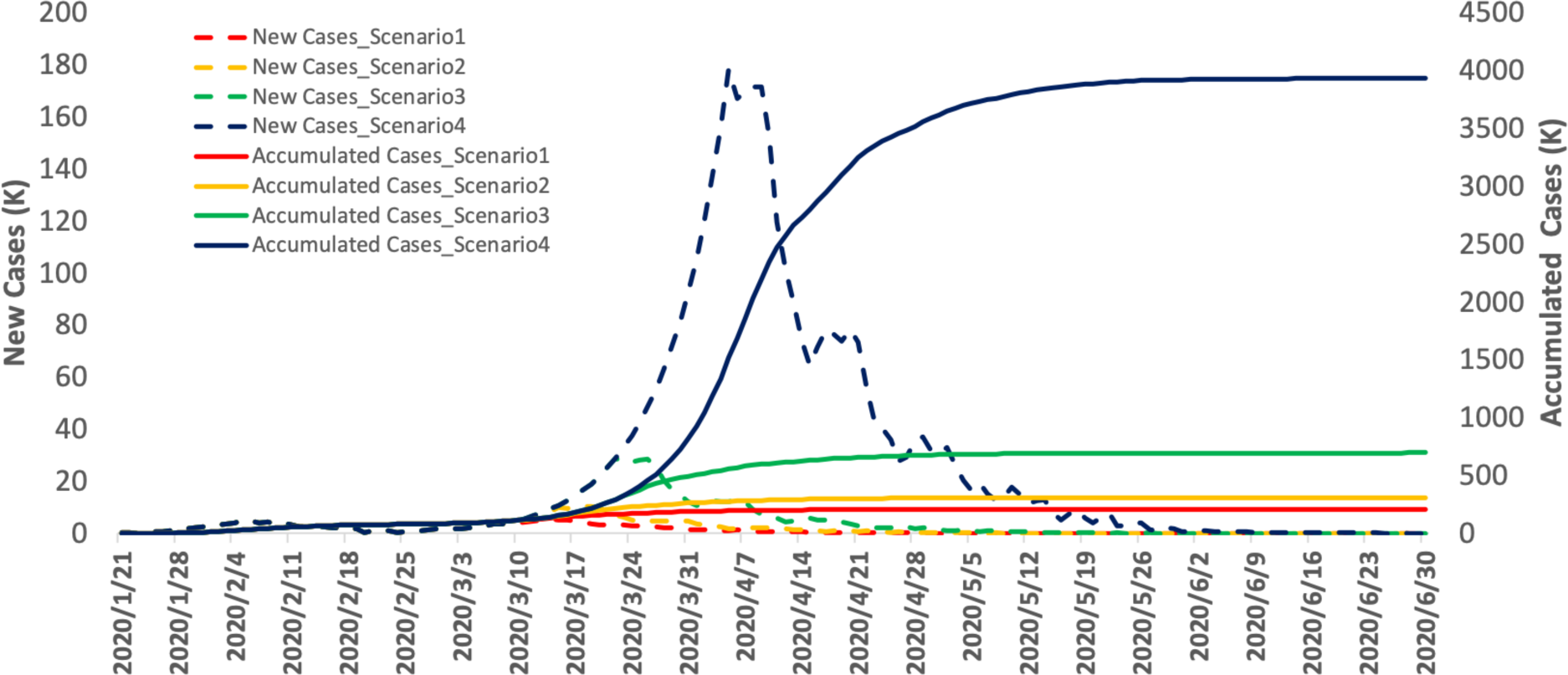
The world reported and forecasted curves of the cumulative and newly confirmed cases of Covid-19 under four intervention scenarios as a function of date from January 21, 2020 to June 18, 2020.

To explore the shape of time-case curves of the top seven infected countries: China, Iran, Italy, Germany, South Korea, USA, France and all other countries, we presented Figure S3. We observed two remarkable features. First, the time-case curve under the later intervention scenario was shifted for more than one month to the right, which implied that both peak time and end time lasted longer. Second, the time-case curve under later intervention was much steeper than that of under the early intervention. In other words, later intervention will substantially increase the number of cumulative cases of COVID-19.

Figure S4 showed the case-fatality rate curve as a function of the Date where the case-fatality rate was defined as the ratio of the number of deaths over the number of cumulative cases in the world. The average case-fatality rate was 3.4%.

## Discussion

As an alternative to the epidemiologic transmission model, we used MAE to forecast the real-time trajectory of the transmission dynamics and generate the real-time forecasts of Covid-19 across the world. The results showed that the accuracies of prediction and subsequently multiple-step forecasting were high. This approach allows us to address two important questions. The first question is whether comprehensive non-drug public health intervention is required or not. The second question is how important is the intervention time. Since interventions are complicated and are difficult to quantify, we designed four intervention scenarios to represent the degrees of interventions and delay of interventions. The proposed methods combine the real data and some assumptions. This allowed us to evaluate the consequences of intervention, while keeping the analysis as close to the real data as possible.

The MAE models allow inputting the interventions information, investigating the impact of interventions on the size, duration and time of the virus outbreak and recommending the intervention time. Using this approach, we also can assess the degree of intervention and group the countries by intervention similarity.

Our analysis demonstrated that public health intervention was extremely important. We observed that delaying intervention for one month caused the maximum number of cumulative cases increase from 211,000 to 3,929,641 and increased 18.62 times. Using 3.4% as the average case fatality rate, the number of deaths increased from 7,174 to 133,608. We analyzed 102 countries and identified 44 countries for which delaying intervention for one month led to serious consequences. For example, delaying intervention for one month in Italy, Iran, USA and Germany increased the maximum number of cumulative cases from 29,475 to 1,493,498, from 37,428 to 729,274, from 5,278 to 293,797, and from 8,795 to 144,542, respectively. We also simulated the impact of delaying intervention for one week or two weeks on controlling the spread of coronavirus (scenarios 2 and 3). The impact of delaying one or two weeks on the spread of the coronavirus was between the immediate intervention and delaying intervention for one month. Intervention will provide the greatest benefit to mitigate the epidemic.

Complimentary to a model approach to transmission dynamics of virus outbreaks, the data driven AI-based methods provide real time forecasting tools for tracking, estimating the trajectory of epidemics, assessing their severity, and predicting the lengths of epidemics. We estimated the duration, peak time and ending time, peak number and maximum number of cumulative cases of Covid-19 under several comprehensive intervention scenarios for 102 countries across the world. This provided important information for government and health workers to make urgent public health response planning to control the spread of Covid-19.

## Data Availability

The very raw data could be found from the WHO website at https://www.who.int/emergencies/diseases/novel-coronavirus-2019/situation-reports. The processed data could be found at https://github.com/wenrurumon/stnn/blob/master/project/data0308.csv and the codes for the analytic framework could be found at https://github.com/wenrurumon/stnn/blob/master/project/.

https://github.com/wenrurumon/stnn/blob/master/project/data0308.csv

https://github.com/wenrurumon/stnn/blob/master/project/

## Supplementary Materials

Material and Methods

Figs. S1 and S2

Table S1, S2 and S3

References (13-16)

## Acknowledgments

**Funding**

### Author contributions

Z,H developed the software and conducted the data analysis. Q,G and S,L conducted the data analysis. M,X designed the project and wrote manuscript, L,J designed the project.

### Data and materials availability

COVID-19 data could be found from WHO at https://www.who.int/emergencies/diseases/novel-coronavirus-2019/situation-reports. The process of data processing and modeling could be fould at https://github.com/wenrurumon/stnn/tree/master/project.

